# Sputum quality and diagnostic outcomes of GeneXpert MTB/RIF assay among adults with presumed tuberculosis in two district hospitals in Ghana: A retrospective cross-sectional study

**DOI:** 10.64898/2026.11.17.26368278

**Authors:** Alhassan Osumanu Hishman, David Taabe, Ofosu Boateng Joseph, Kwabena Adade Kusi, Felix Kodzo Besah Sorvor, Evans Ansu Twumasi, Akosua Frimpomaa Amoateng, Johnpaul Amenu, Osei Kwame Wiredu, Emmanuel Boateng Mensah, Antoinette Salim Saaiby, Baffour Osei Akoto, Edward Owusu Kwarteng, Michael Cappello, Peter Kojo Brenya

## Abstract

**Background:** Tuberculosis remains a major public health challenge in Ghana, where GeneXpert MTB/RIF serves as a first–line diagnostic tool. However, uncertainties about this method persist regarding specimen quality and the clinical significance of indeterminate rifampicin susceptibility results.

**Methods:** We conducted a retrospective cross–sectional study of samples from 3,977 presumptive TB patients tested either at HopeXchange Medical Center (Kumasi) or Keta Municipal Hospital between January 2020 and December 2024. Demographic data, sputum macroscopic appearance, and GeneXpert results were analyzed using descriptive statistics and Fisher’s exact test.

**Results:** Overall *Mycobacterium tuberculosis* (MTb) test positivity was 11.6% (461/3977). Males accounted for 63.3% of positive results, with the highest percentage among those ages 30–40 years. Mucopurulent specimens yielded the highest positivity rate (25.4%), compared to 4.0% of salivary specimens. Blood–stained sputum specimens (n=44) showed a 15.9% positivity rate. Although confirmed rifampicin resistance was low (1.7%), 9.3% of all MTb positive samples yielded an indeterminate result. The rate of indeterminate results from samples collected at Kwadaso Municipal increased from 7.3% in 2021 to 28.6% in 2024.

**Conclusion:** These findings validate Ghana National Tuberculosis Program (NTP) guidelines emphasizing the reliability of mucopurulent specimen collection, but challenges the routine rejection of blood–stained sputum. The rising trend of indeterminate rifampicin results requires further investigation of circulating *rpoB* mutation profiles and quality assurance measures. Patients with indeterminate results should be further assessed using drug susceptibility testing to rule out rifampicin resistance. The findings provide actionable evidence to refine diagnostic algorithms and improve TB care in Ghana.

## INTRODUCTION

Tuberculosis (TB), an infection caused by *Mycobacterium tuberculosis* (MTb) (1), remains a major threat to global public health. Despite efforts in prevention and treatment, this disease continues to cause significant worldwide burden of morbidity and mortality especially in low or limited resource countries. A 2024 World Health Organization Global Tuberculosis Report affirms the persistent nature of this epidemic, highlighting over a million deaths and millions of new cases annually (2). African countries bear a disproportionate share of this burden due to challenges in healthcare infrastructure and lack of access to advanced diagnostics.

Surveillance for TB constitutes a major health priority in Ghana. In the same 2024 WHO report, an estimated 44,000 individuals developed TB in the country, with approximately 9,900 deaths attributable to the disease in just a single year(2,3). This burden of TB mortality highlights persistent challenges across the TB diagnostic and care pathway, particularly in achieving early and accurate diagnosis, timely treatment initiation, and effective case management. Efficient and effective TB control requires rapid identification and isolation of infected individuals in order to interrupt transmission, a goal that is fundamentally dependent on reliable diagnostic systems. For decades, TB diagnosis relied heavily on smear microscopy(4), a method with limited sensitivity, and culture, which is slow and requires highly skilled technical staff.

The introduction of the GeneXpert MTB/RIF platform, an automated, cartridge-based nucleic acid amplification test (NAAT), has reshaped TB diagnosis in high-burden, resource-limited settings like Ghana(5). The WHO-approved technology is able to concurrently detect both MTb complex DNA and rifampicin resistance mutations within two hours(6). This approach offers diagnostic improvement in speed and sensitivity compared to smear microscopy methods(7). In Ghana, the National Tuberculosis Control Programme (NTP) together with the Ghana Health Service (GHS) has approved GeneXpert MTB/RIF as the first line diagnostic test for all presumptive pulmonary TB cases. The expansion of GeneXpert MTB/RIF has been associated with improved TB case finding and facilitated the linkage of diagnosed patients to treatment (8).

The analytical performance of this molecular tool is dependent on the quality of the sputum specimen. Sputum remains as the preferred primary specimen for pulmonary TB laboratory investigations, and its macroscopic quality directly influences detection sensitivity(9). NTP classifies sputum into four categories based on appearance: mucopurulent (purulent, viscous); muco-salivary (a mixture); salivary (watery or clear saliva); and blood–stained. Deep-cough, early-morning, mucopurulent sputum is universally considered the optimal sample. This may, however, be affected by the greater proportion of suboptimal specimens collected in low-resource health facilities with clinical and logistic constraints.

A common practice in many laboratories is the routine rejection of blood-stained sputum samples submitted for GeneXpert testing. This practice most often rests on the concern that heme compounds in blood may inhibit polymerase chain reaction (PCR) activity, ultimately leading to false-negative MTb results and/or inaccurate rifampicin resistance profiles(10,11). These routine operational principles are often based on anecdotal reports in the absence of rigorously acquired evidence, thereby leading to additional diagnostic delay that negatively impacts quality of care. For example, patients presenting with symptoms of hemoptysis, a condition associated with advanced pulmonary pathology such as cavitary disease may experience diagnostic delays when their blood–stained sputum sample is rejected, postponing lifesaving treatment and potentially facilitating ongoing transmission.

The evidence base underlying this practice is inconclusive, creating a gap between operational principles and sound scientific data. However, recent reports suggest that the practice of rejecting blood-stained specimens may not be necessary. Sinshaw et al. (12) demonstrated a high-rate of MTb positivity in blood–stained sputum specimens, confirming a high bacillary burden in these samples. Aninagyei and colleagues (13) investigated the effect of sputum stained with blood on PCR, finding that the GeneXpert assay was reliable at blood contamination level of less than 2%. Levels of blood contamination above 5% reduced the reliability of the assay, with complete PCR inhibition occurring at 20% contamination.

Given the high TB burden in Ghana, dependance on GeneXpert technology, and the unresolved operational challenges around specimen suitability, generating evidence of assay performance will inform diagnostic practices and public health policy. The study described here was undertaken in order to evaluate the diagnostic yield of GeneXpert MTB/RIF assay using the NTP’s sputum quality categories (mucopurulent, muco–salivary, salivary and blood stained) collected from presumptive TB patients in 2 Ghana healthcare settings. Findings from this retrospective study provide new evidence intended to inform diagnostic algorithms, improve laboratory efficiency and ensure that no patient is denied a timely TB diagnosis.

## METHODS

### Ethical considerations and review

Approval for this study was granted by the University for Development Studies Institutional Review Board (USD IRB) (reference number: UDS/ R13/0025/26) and HopeXchange Institutional Review Committee (reference number: HXCMC/GL/VOL1/2025/256).

### Study design and site

This retrospective cross-sectional study design included TB registers from two TB testing hubs in separate regions of Ghana (Fig 1). The registers contained records of patients who underwent TB testing, including demographic characteristics and laboratory test results. HopeXchange Medical Center (HXCMC) is a Catholic Mission Hospital located in the Kwadaso Municipality, Kumasi, Ashanti Region. The HXCMC laboratory, which is accredited as a testing hub by the Ghana National Tuberculosis Program (NTP), operates two GeneXpert analyzers, each of which has 4 modules for testing. Within the Municipality, 32 health facilities were assigned as sites for collection of sputum specimens (spokes), which were transported to HXCMC medical microbiology laboratory for testing. The second study site was Keta Municipal Hospital (KMH), which is government hospital found in the Volta Region. KMH laboratory is also accredited by NTP as a TB testing hub and serves 6 spoke health facilities. The KMH operates one GeneXpert analyzer with 4 modules for testing.

**Figure 1:**
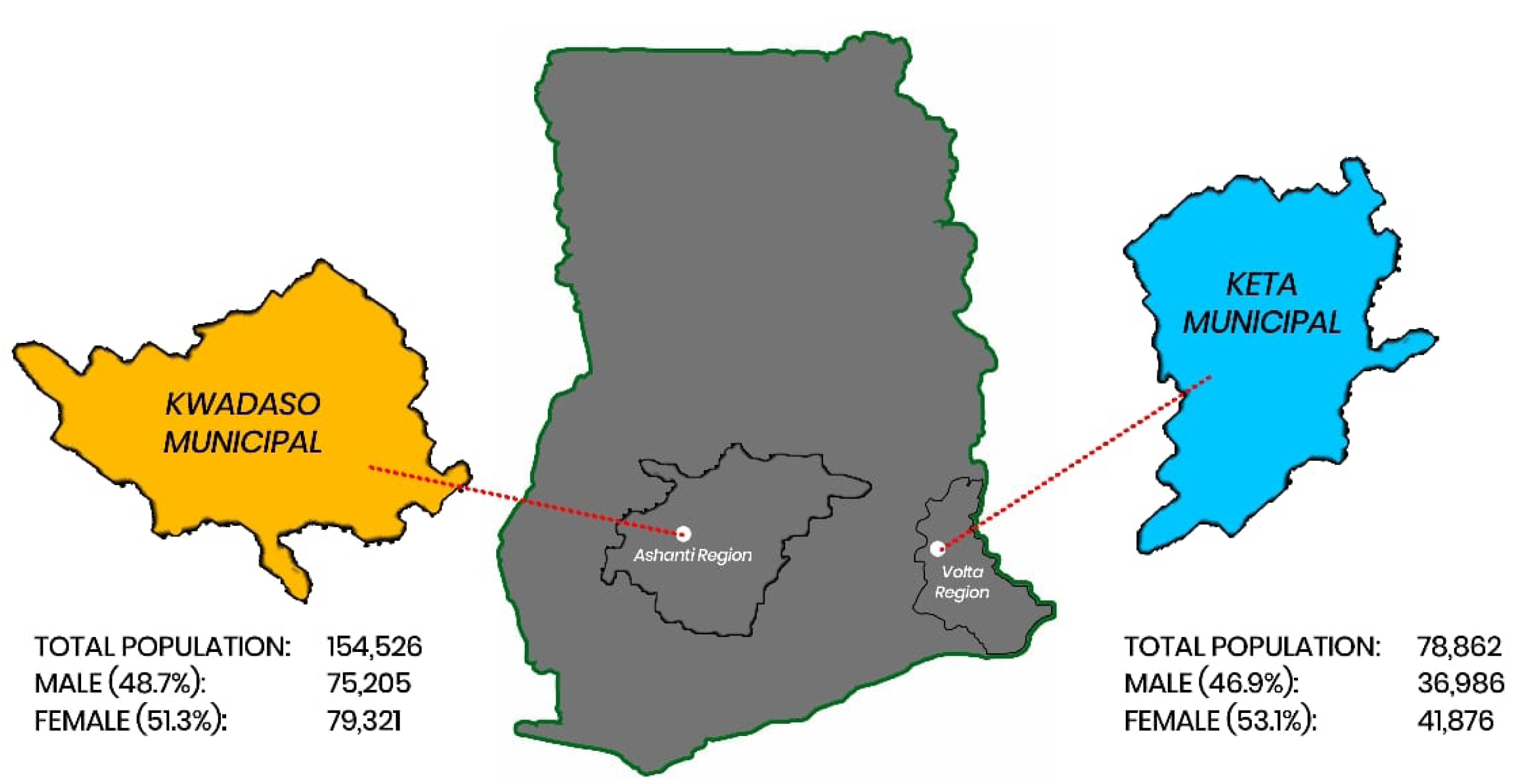
Map of selected facilities

**Figure 2:**
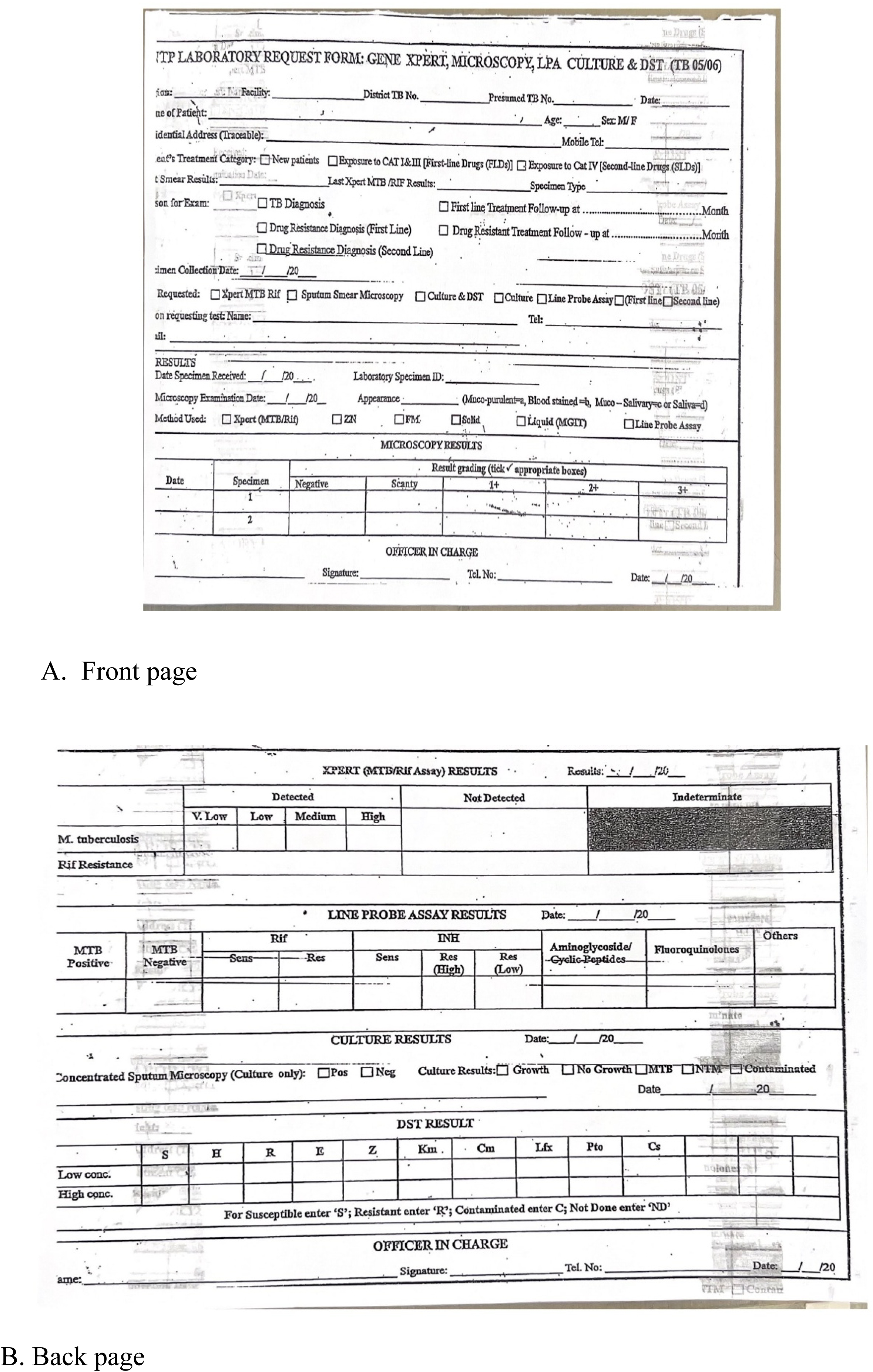
Sample Tb request form

### Study population

Records of presumed TB patients who underwent initial testing by GeneXpert MTB/RIF assay between January 2020 and December 2024 were reviewed for the study. Adults who were 18 years of age with no prior TB testing history were included in the study. Records with missing demographic or specimen data, or with sputum volume < 1mL were excluded, as were duplicate records. In total, 3,977 records met the inclusion criteria and were considered for analysis.

### Data collection and analysis

Information on gender, age, sputum appearance and results of GeneXpert were retrieved from the TB laboratory registers and entered into a Microsoft Excel spreadsheet. Data analysis was performed using IBM SPSS version 27. Descriptive statistics and Fischer exact test were applied and a p–value < 0.05 was considered statistically significant.

## RESULTS

A total of 3,977 records were retrieved from two Ghanaian TB Hub site registers for TB case finding and resistance monitoring. As shown in Table 1, there was a statistically significant association between GeneXpert MTb testing result and age, gender, and sputum appearance. The highest MTb detection rate (25.8%; n=119) was found in patients between 30 and 49 years of age. Male patients accounted for 63.3% of all positive results, despite making up less than half (42.2%) of the study population tested. The physical characteristic of the sputum sample was also a strong predictor of GeneExpert result, with samples described as mucopurulent showing the highest percentage of positive tests (26.0%) compared to muco-salivary, salivary and blood stained. The majority of the samples (3112/3977; 78.2%) were collected from subjects at HopeXchange Medical Center (Kwadaso Municipality), with a GeneXpert MTb positive rate of 12.3% (n=384). By comparison, 77 of the 865 (8.9%) samples collected at Keta Municipal Hospital were positive for MTb.

**Table 1.** : Statistical association between Socio-Demographics and Sputum Macroscopic Appearance with GeneXpert MTB/RIF Positivity.

| **Characteristic** | **Study Cohort (N=3977)** | **MTb Positive (n=461)** | **MTb Negative (n=3516)** | **□2 (p-value)** |
|---|---|---|---|---|
| Age, Years (Mean ± SD)* | 45.0 ± 16.6 | 43.4 ± 13.8 | 45.3 ± 16.9 | 0.009 |
| **Age Category** |  |  |  | 27.85 (<0.001) |
| 18 - 29 years | 817 | 76 (9.3%) | 741 (90.7%) |  |
| 30 - 39 years | 824 | 119 (14.4%) | 705 (85.6%) |  |
| 40 - 49 years | 828 | 119 (14.4%) | 709 (85.6%) |  |
| 50 - 59 years | 687 | 82 (11.9%) | 605 (88.1%) |  |
| ≥ 60 years | 821 | 65 (7.9%) | 756 (92.1%) |  |
| **Gender** |  |  |  | 61.33 (< 0.001) |
| Male | 1839 | 292 (15.9%) | 1547 (84.1%) |  |
| Female | 2138 | 169 (7.9%) | 1969 (92.1%) |  |
|  | ***Total tested, n*** | ***Positive, n (%)*** | ***Negative, n (%)*** |  |
| **Sample Appearance** |  |  |  | 129.29 (< 0.001) |
| Mucopurulent | 473 | 120 (25.4) | 353 (74.6) |  |
| Salivary | 721 | 29 (4.0) | 692 (96.0) |  |
| Muco-salivary | 2739 | 305 (11.1) | 2434 (88.9) |  |
| Blood - Stained | 44 | 7 (15.9) | 37 (84.1) |  |
| **Total** | 3977 | 461 (11.6) | 3516 (88.4) |  |
| **Study Site** |  |  |  | 7.81 (0.005) |
| KMH | 865 | 77 (8.9) | 788 (91.1) |  |
| HXCMC | 3112 | 384 (12.3) | 2728 (87.7) |  |

\* mean ± standard deviation using T-test

Among the 461 MTb positive samples from both study locations, there were 7 (1.5%) that exhibited rifampicin resistance mutations (Table 2). However, 43/461 (9.3%) of all MTb positive samples showed an indeterminate result regarding rifampicin resistance. The rate of rifampicin resistance observed among MTb positive samples collected across the study period (2021–2024) varied from 0-2.07%.

**Table 2:**
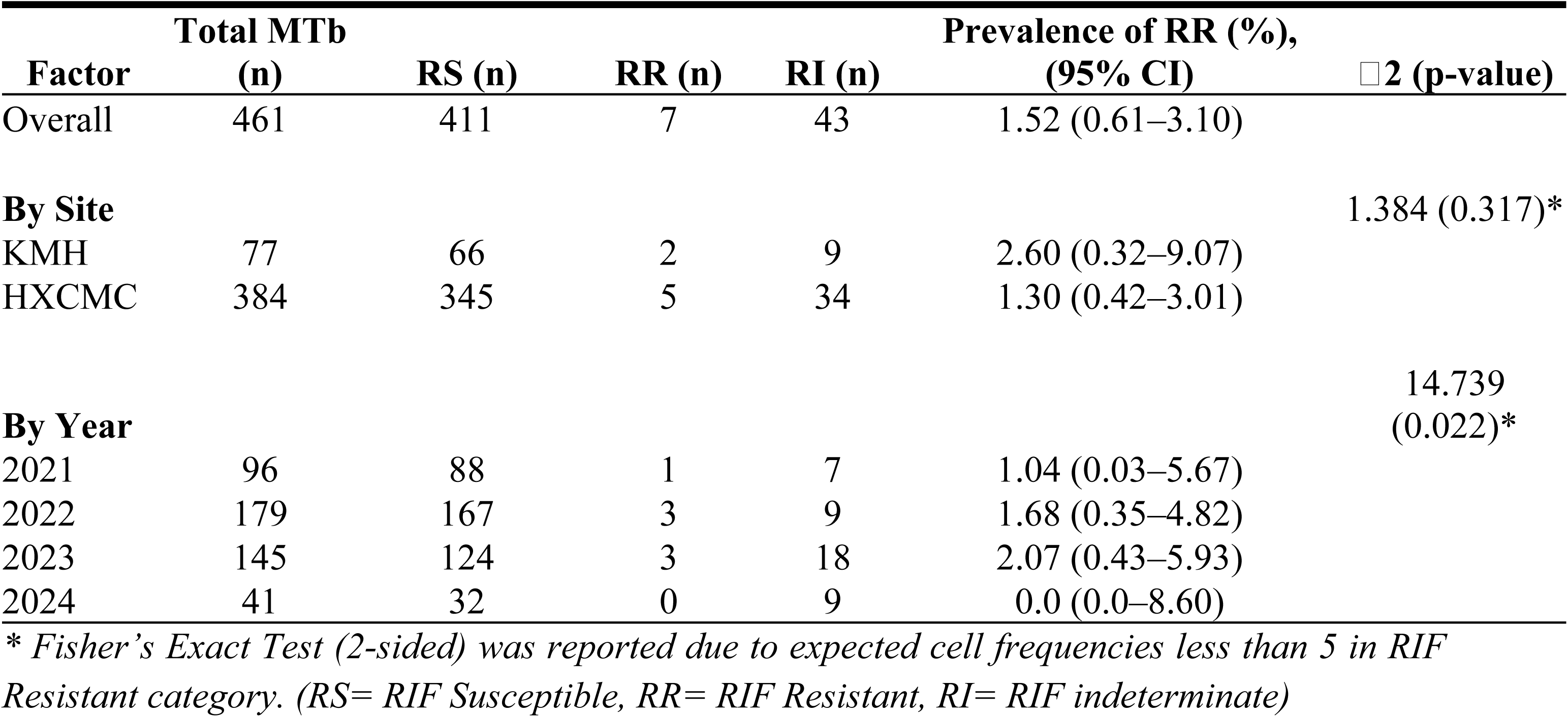
Epidemiological Distribution of Rifampicin Resistance and Indeterminate Results by Clinical Site and Temporal Markers.

As shown in Table 3, KMH samples revealed a consistent MTb positivity rate (10.8%) in 2022 – 2023, followed by a decrease to 5.9% in 2024. By contrast, the rate of MTb positivity at HXCMC revealed a downward trend from 2021 (16.6%) to 2024 (9.2%). Table 3 shows a consolidated annual comparison between KMH and HXCMC and the emerging issues of increasing indeterminate results over time.

**Table 3:** Comparative Annual Inventory of GeneXpert MTB/RIF Diagnostic Outcomes and Drug Susceptibility Across Ghanaian District Hubs.

| **Year/Hospital** | **Total Test Conducted (N)** | **MTB Positive, n (%)** | **P-value (MTb Positive)** | **RR among Positives, n (%)** | **P-value (RIF Resistance)** | **RIF susceptible (RS), n (%)** | **RIF Resistant (RR), n (%)** | **RIF Indeterminate (RI), n (%)** |
|---|---|---|---|---|---|---|---|---|
| **HXCMC** |  |  |  |  |  |  |  |  |
| 2020 | 20 | 0 (0.0) | (0.001) * | 0 (0.0) | (< 0.001) * | 0 (0.0) | 0 (0.0) | 0 (0.0) |
| 2021 | 579 | 96 (16.6) |  | 1 (1.0) |  | 88 (91.7) | 1 (1.0) | 7 (7.3) |
| 2022 | 1183 | 152 (12.8) |  | 3 (1.7) |  | 144 (94.7) | 2 (1.3) | 6 (4.0) |
| 2023 | 1095 | 115 (10.5) |  | 3 (2.1) |  | 98 (85.2) | 2 (1.7) | 15 (13.1) |
| 2024 | 228 | 21 (9.2) |  | 0 (0.0) |  | 15 (71.4) | 0 (0.0) | 6 (28.6) |
| **KMH** |  |  |  |  |  |  |  |  |
| 2022 | 250 | 27 (10.8) |  | - |  | 23 (85.2) | 1 (3.7) | 3 (11.1) |
| 2023 | 278 | 30 (10.8) |  | - |  | 26 (86.7) | 1 (3.3) | 3 (10.0) |
| 2024 | 337 | 20 (5.9) |  | - |  | 17 (85.0) | 0 (0.0) | 3 (15.0) |
| Total | 3977 | 461 (11.6) |  | - |  | 411 (89.2) | 7 (1.5) | 43 (9.3) |

\* *P-value for trend calculated using Fisher’s Exact Test(2-sided)*

## DISCUSSION

Retrospective data analysis of 3,977 clinical samples from patients suspected of MTb infection demonstrated GeneXpert MTB/RIF detection of *Mycobacterium tuberculosis* in all sputum categories, including blood – stained specimens. Given the importance of timely diagnosis of active tuberculosis in Ghana and other endemic populations, the data presented here suggests that the common laboratory practice of rejecting specimens based on perceived quality needs to be revisited. Nonetheless, the finding that mucopurulent sputum specimens were more than twice as likely to test positive than other sputum types affirms the value of obtaining high-quality specimens whenever possible. Moreover, we demonstrate the value of applying molecular diagnostic techniques in endemic settings using locally acquired samples obtained from patients.

The higher rate (25.4%) of TB detection observed in mucopurulent sputum specimens is consistent with accepted understanding of pulmonary physiology in the setting of bacterial infection. Mucopurulent sputum originates from the lower respiratory tract, often a site of active infection and inflammation, and therefore would be expected to harbor a higher bacillary load. This finding validates the Ghana NTP guidelines, which promotes collection of deep – cough specimens when TB is suspected. Suboptimal salivary specimens, which may consist mostly of upper respiratory secretions, revealed poor diagnostic yield in our study, with a positivity rate of 4.0%. Such a clear difference in yield confirms that effective patient instruction and quality specimen collection are important first steps in the diagnostic cascade. Medical Laboratory Professionals and Physicians must reinforce this message, as the technological capacity of GeneXpert cannot compensate for poor – quality samples (14–16).

Common practice in Ghana and other low-resource settings is to recommend routine rejection of blood–stained sputum specimens over fears of PCR inhibition. However, the manufacturer’s (Cepheid) instructions for use state that the proprietary sample reagent is designed to lyse cells, inactivate *M. tuberculosis*, and neutralize common PCR inhibitors, including heme(17). Data from this study support these guidelines, as the data showed a MTb positivity rate of 15.9% (7/44) in blood-stained samples. A study carried out in Ethiopia also reported similar high detection rate in blood stained sputum specimens (18). Hemoptysis, in clinical practice, is a serious pathological condition linked with advanced cavitary TB (19–22). Deferring presumed TB patients for fresh collection of specimens free from blood before processing creates potentially dangerous diagnostic delays, leading to postponement of treatment for individuals with high suspicion of active infection. Our results strongly argue for a change in the policy of rejecting blood-stained samples, which instead should be considered suitable for processing and analysis by GeneExpert.

The demographic distribution of positive cases, with significant predominance among males (63.3%) and individuals aged 30 – 49 years reflects an epidemiologic pattern commonly reported in high – burden settings(23). A potentially higher risk among men may be linked to social, occupational, and healthcare – seeking behaviors, as noted in the WHO’s 2024 Global TB report, which highlights gender disparities in case notification. The disproportionate burden in the economically productive age group underscores TB’s substantial social and economic impact, justifying the need for sustained public health investment.

The data presented here support the Ghana NTP’s current recommendations regarding first-line TB therapy, given the low prevalence of confirmed rifampicin resistance (1.7%) within the study population. Of note, this figure is below the 2024 national estimate of TB incidence (126 cases per 100,000 population) reported in the latest WHO 2025 global report, which may reflect regional variation(24,25). This observation, however, is somewhat confounded by the fact that 9.3% of TB positive samples exhibited indeterminate rifampicin resistance by GeneExpert. An indeterminate rifampicin resistance result does not necessarily represent a test failure, but rather indicates that the assay could not definitively classify the resistance status based on DNA mutation sequences pre–programmed in the GeneXpert analyzer(26–28). The assay uses five overlapping probes (A through E) to interrogate the 81 – base pair rifampicin – resistance – determining region (RRDR) of the *rpoB* gene(29). Rifampicin resistance is determined from changes in the probe signals generated across this region, and atypical probes signals may result in rifampicin – resistance results that require further investigation. (30)

This is of potential clinical significance, as novel *rpoB* mutations can produce atypical probe – binding or melting-temperature patterns that may not be readily interpreted by the assay algorithm. Experimental studies with Xpert MTB/RIF ultra have demonstrated that certain mutation combinations can results in an indeterminate rifampicin–resistance call when the melting – temperature profile falls outside the software’s predefined detection window (31).

Our findings of 9.3% positives indeterminate rifampicin exceed 0.7% to 1.2% range reported in studies from the Philippines, India, and Ethiopia (32,33). The clinical significance of indeterminate results is unclear, as confirmation by culture has identified both susceptible and resistant specimens (34). However, the practical consequence of an indeterminate result for rifampicin resistance creates genuine clinical ambiguity. Patients diagnosed with TB and an indeterminate resistance profile may need to wait weeks for phenotypic culture results, causing critical delays in initiating appropriate therapy. By contrast, empirically treating all indeterminate results as resistant would expose many patients unnecessarily to toxic second–line drugs.

Rigouts and colleagues demonstrated that mutations at codon 432 and the Lys446Gln mutation consistently yield indeterminate results on GeneXpert Ultra (35). They concluded that indeterminate results “may indicate underlying *rpoB* mutations within the rifampicin – resistance determining region and thus increase the post – test probability of rifampicin resistance, albeit to an unknown extent” (35). Naik and colleagues also compared 538 trace – positive, rifampicin – indeterminate specimens against phenotypic MGIT drug susceptibility testing and found that 16.2% were true rifampicin – resistant (36). A similar study comparing 31 rifampicin – indeterminate Xpert MTB/ RIF Ultra results with phenotypic culture and drug susceptibility testing found 2 cases (6.5%) confirming rifampicin resistant. Conversely, a study in Uganda found no missed resistant among 24 cultured isolates, highlighting that predictive value varies by setting and circulating strains(37).

An increase in the proportion of indeterminate rifampicin results was observed in samples from HopeXchange Medical Center across the retrospective study period, from 7.3% in 2021 to 28.6% in 2024. If these results are determined to be associated with rifampicin resistance, then careful follow-up is necessary given that HopeXchange serves as a TB Hub within the Kwadaso Municipality. Temporal shifts in predominant strain lineages or novel resistance mutations may be present in the communities surveyed, potentially altering probe-binding patterns leading to an increase in indeterminate results (38). Systematic characterization of these mutations through targeted next – generation sequencing or line – probe assays could potentially clarify whether the trend reflect changing molecular epidemiology or operational factor(39,40).

## Conclusion

The findings reported here validate Ghana National Tuberculosis Program (NTP) guidelines emphasizing mucopurulent specimen collection, while challenging the routine rejection of blood – stained sputum, which can yield clinically meaningful rates of TB detection. The rising trend of indeterminate rifampicin resistance results requires an urgent investigation into circulating *rpoB* mutation profiles and quality assurance measures, especially Kwadaso Municipality. Patients with indeterminate results require careful management and in vitro drug susceptibility testing, as these may indicate rifampicin resistance. Clinicians need education to raise awareness that indeterminate results carry a non–negligible probability of rifampicin resistance, necessitating prompt clinical assessment and institution of enhanced infection control precautions. These findings provide actionable evidence to refine diagnostic algorithms and improve TB care in Ghana

## Acknowledgements

The authors would like to thank the clinical laboratory staff, healthcare providers, administrators and patients at HopeXchange Medical Centre and Keta Municipal Hospital for their participation and contributions to the study.

## Data Availability

Raw data used to support findings of this study have been deposited in the Zenodo open repository with the digital object identifier (DOI) 10.5281/zenodo.24146151.

## REFERENCES

[1] A. Maciejak-Jastrzębska, G. Sygitowicz, S. Brzezińska, K. Bielska, and E. Augustynowicz-Kopeć, “Tuberculosis Today: Microbial Insights, Epidemiological Trends, and the Role of Molecular Diagnostics,” Pathogens, vol. 14, no. 10, p. 965, Oct. 2025, doi: 10.3390/pathogens14100965.

[2] World Health Organization, “Global Tuberculosis Report 2024. Geneva: WHO; 2024. Licence: CC BY-NC-SA 3.0 IGO.,” Global tuberculosis report, p. 68, 2024, Accessed: Mar. 26, 2026. [Online]. Available: https://www.who.int/publications/i/item/9789240101531

[3] F. Estaji, A. Kamali, and M. Keikha, “Strengthening the global Response to Tuberculosis: Insights from the 2024 WHO global TB report,” J. Clin. Tuberc. Other Mycobact. Dis., vol. 39, May 2025, doi: 10.1016/j.jctube.2025.100522.

[4] N. Zaporojan, R. A. Negrean, R. Hodișan, C. Zaporojan, A. Csep, and D. C. Zaha, “Evolution of Laboratory Diagnosis of Tuberculosis,” Clin. Pract., vol. 14, no. 2, pp. 388–416, Apr. 2024, doi: 10.3390/clinpract14020030.

[5] J. K. Boakye-Appiah et al., “High prevalence of multidrug-resistant tuberculosis among patients with rifampicin resistance using GeneXpert Mycobacterium tuberculosis/rifampicin in Ghana,” Int. J. Mycobacteriol., vol. 5, no. 2, pp. 226–230, 2016, doi: 10.1016/j.ijmyco.2016.02.004.

[6] K. Weyer et al., “Rapid molecular TB diagnosis: evidence, policy making and global implementation of Xpert MTB/RIF,” European Respiratory Journal, vol. 42, no. 1, pp. 252–271, Jun. 2013, doi: 10.1183/09031936.00157212.

[7] K. K. Asare et al., “Comparison of microscopic and xpert MTB diagnoses of presumptive mycobacteria tuberculosis infection: retrospective analysis of routine diagnosis at Cape Coast Teaching Hospital,” BMC Infect. Dis., vol. 24, no. 1, Dec. 2024, doi: 10.1186/s12879-024-09566-9.

[8] J. K. Boakye-Appiah et al., “High prevalence of multidrug-resistant tuberculosis among patients with rifampicin resistance using GeneXpert Mycobacterium tuberculosis/rifampicin in Ghana,” Int. J. Mycobacteriol., vol. 5, no. 2, pp. 226–230, Jun. 2016, doi: 10.1016/J.IJMYCO.2016.02.004.

[9] J. E. Coronel et al., “Evaluation of microbiological variants of sputum processing and concentration of mycobacteria to optimize the microscopic and imaging diagnosis of tuberculosis,” Int. J. Mycobacteriol., vol. 8, no. 1, pp. 75–82, Jan. 2019, doi: 10.4103/IJMY.IJMY_172_18.

[10] A. J. Meyer et al., “Sputum quality and diagnostic performance of GeneXpert MTB/RIF among smear-negative adults with presumed tuberculosis in Uganda,” PLoS One, vol. 12, no. 7, p. e0180572, Jul. 2017, doi: 10.1371/JOURNAL.PONE.0180572.

[11] N. Taylor, R. L. Gaur, E. J. Baron, and N. Banaei, “Can a Simple Flotation Method Lower the Limit of Detection of Mycobacterium tuberculosis in Extrapulmonary Samples Analyzed by the GeneXpert MTB/RIF Assay?,” J. Clin. Microbiol., vol. 50, no. 7, p. 2272, Jul. 2012, doi: 10.1128/JCM.01012-12.

[12] W. Sinshaw et al., “Effect of sputum quality and role of Xpert® MTB/ RIF assay for detection of smear-negative pulmonary tuberculosis in same-day diagnosis strategy in Addis Ababa, Ethiopia,” Afr. J. Lab. Med., vol. 11, no. 1, p. 1671, 2022, doi: 10.4102/AJLM.V11I1.1671.

[13] E. Aninagyei, R. Ayivor-Djanie, J. Attoh, M. P. Dakorah, M. N. Ginko, and D. O. Acheampong, “Molecular detection of Mycobacterium tuberculosis in blood stained sputum samples using GeneXpert PCR assay,” Diagn. Microbiol. Infect. Dis., vol. 100, no. 3, Jul. 2021, doi: 10.1016/J.DIAGMICROBIO.2021.115363.

[14] D. A. Geleta, Y. C. Megerssa, A. N. Gudeta, G. T. Akalu, M. T. Debele, and K. D. Tulu, “Xpert MTB/RIF assay for diagnosis of pulmonary tuberculosis in sputum specimens in remote health care facility,” BMC Microbiology 2015 15:1, vol. 15, no. 1, pp. 220-, Oct. 2015, doi: 10.1186/S12866-015-0566-6.

[15] A. J. Meyer et al., “Sputum quality and diagnostic performance of GeneXpert MTB/RIF among smear-negative adults with presumed tuberculosis in Uganda,” PLoS One, vol. 12, no. 7, p. e0180572, Jul. 2017, doi: 10.1371/JOURNAL.PONE.0180572.

[16] C. M. Denkinger et al., “Defining the Needs for Next Generation Assays for Tuberculosis,” J. Infect. Dis., vol. 211, no. suppl_2, pp. S29–S38, Apr. 2015, doi: 10.1093/INFDIS/JIU821.

[17] “Operator Manual GeneXpert ® System with Touchscreen”.

[18] W. Sinshaw et al., “Effect of sputum quality and role of Xpert® MTB/RIF assay for detection of smear-negative pulmonary tuberculosis in same-day diagnosis strategy in Addis Ababa, Ethiopia,” Afr. J. Lab. Med., vol. 11, no. 1, p. 7, Aug. 2022, doi: 10.4102/AJLM.V11I1.1671.

[19] S. Iqbal, M. Nassar, R. Kondaveeti, T. O’Brien, R. S. Siddiqui, and R. Lopez, “Massive Hemoptysis in Previously Treated Pulmonary Tuberculosis: A Case Series,” Cureus, vol. 14, no. 8, p. e27882, Aug. 2022, doi: 10.7759/CUREUS.27882.

[20] J. R. Middleton, P. Sen, M. Lange, J. Salaki, R. Kapila, and D. B. Louria, “Death-Producing Hemoptysis in Tuberculosis,” Chest, vol. 72, no. 5, pp. 601–604, Nov. 1977, doi: 10.1378/CHEST.72.5.601.

[21] L. van de Sand, B. M. Schaarschmidt, J. Wienker, O. Witzke, and M. Zettler, “Hemoptysis in a patient with MDR-tuberculosis: successful diagnosis with photon counting CT and embolization of a Rasmussen aneurysm,” Infection, vol. 54, no. 2, p. 983, Apr. 2025, doi: 10.1007/S15010-025-02660-3.

[22] V. A. Brady, A. V. Zinchuk, M. D. Siegel, and J. D. Possick, “A Young Man With Hemoptysis and Cavitary Lung Lesions,” Chest, vol. 153, no. 4, p. e85, Apr. 2018, doi: 10.1016/J.CHEST.2017.09.018.

[23] World Health Organization, “Global Tuberculosis Report 2024. Geneva: WHO; 2024. Licence: CC BY-NC-SA 3.0 IGO.,” Global tuberculosis report, p. 68, 2024, Accessed: Apr. 30, 2026. [Online]. Available: https://www.who.int/publications/i/item/9789240101531

[24] World Health Organization, “Global tuberculosis report 2025,” Geneva, 2025.

[25] H. K. Hackman et al., “Treatment outcomes among tuberculosis patients with co-morbidities at Ghana’s premier national referral centre: a retrospective cross-sectional study,” BMC Infectious Diseases 2026 26:1, vol. 26, no. 1, pp. 608-, Feb. 2026, doi: 10.1186/S12879-026-12884-9.

[26] L. Rigouts, J. Keysers, R. Rabab, K. Fissette, A. van Deun, and B. C. de Jong, “GeneXpert MTB/RIF Ultra performance to detect uncommon rpoB mutations in Mycobacterium tuberculosis,” BMC Research Notes 2023 16:1, vol. 16, no. 1, pp. 146-, Jul. 2023, doi: 10.1186/S13104-023-06394-Z.

[27] C. C. Boehme et al., “Feasibility, diagnostic accuracy, and effectiveness of decentralised use of the Xpert MTB/RIF test for diagnosis of tuberculosis and multidrug resistance: A multicentre implementation study,” The Lancet, vol. 377, no. 9776, pp. 1495–1505, 2011, doi: 10.1016/S0140-6736(11)60438-8.

[28] A. M. Elbrolosy, R. H. El Helbawy, O. M. Mansour, and R. A. Latif, “Diagnostic utility of GeneXpert MTB/RIF assay versus conventional methods for diagnosis of pulmonary and extra-pulmonary tuberculosis,” BMC Microbiology 2021 21:1, vol. 21, no. 1, pp. 144-, May 2021, doi: 10.1186/S12866-021-02210-5.

[29] G. Mboowa, C. Namaganda, and W. Ssengooba, “Rifampicin resistance mutations in the 81 bp RRDR of rpoB gene in Mycobacterium tuberculosis clinical isolates using Xpert®MTB/RIF in Kampala, Uganda: a retrospective study,” BMC Infect. Dis., vol. 14, no. 1, p. 481, Sep. 2014, doi: 10.1186/1471-2334-14-481.

[30] A. Alemu et al., “Does Xpert® MTB/RIF assay give rifampicin resistance results without identified mutation? Review of cases from Addis Ababa, Ethiopia,” BMC Infect. Dis., vol. 20, no. 1, Jan. 2020, doi: 10.1186/S12879-020-4817-2.

[31] E. A. Ochang et al., “Evaluation of rifampicin resistance and 81-bp rifampicin resistant determinant region of rpoB gene mutations of Mycobacterium tuberculosis detected with XpertMTB/Rif in Cross River State, Nigeria,” Int. J. Mycobacteriol., vol. 5, pp. S145–S146, Dec. 2016, doi: 10.1016/J.IJMYCO.2016.09.007.

[32] T. A. Demissie and D. Belayneh, “Magnitude of Mycobacterium tuberculosis Infection and Its Resistance to Rifampicin Using Xpert-MTB/RIF Assay Among Presumptive Tuberculosis Patients at Motta General Hospital, Northwest Ethiopia,” Infect. Drug Resist., vol. 14, pp. 1335–1341, 2021, doi: 10.2147/IDR.S300585.

[33] R. Reddy and G. Alvarez-Uria, “Molecular Epidemiology of Rifampicin Resistance in Mycobacterium tuberculosis Using the GeneXpert MTB/RIF Assay from a Rural Setting in India,” J. Pathog., vol. 2017, pp. 1–5, 2017, doi: 10.1155/2017/6738095.

[34] B. G. Tenorio, D. J. Uayan, K. F. Lumunsad, A. Estuart, J. C. R. Javier, and C. L. Abad, “The clinical profile of patients with indeterminate Xpert rifampin resistance results in a single tertiary level institution,” Diagn. Microbiol. Infect. Dis., vol. 113, no. 4, p. 117008, Dec. 2025, doi: 10.1016/J.DIAGMICROBIO.2025.117008.

[35] L. Rigouts, J. Keysers, R. Rabab, K. Fissette, A. van Deun, and B. C. de Jong, “GeneXpert MTB/RIF Ultra performance to detect uncommon rpoB mutations in Mycobacterium tuberculosis,” BMC Research Notes 2023 16:1, vol. 16, no. 1, pp. 146-, Jul. 2023, doi: 10.1186/S13104-023-06394-Z.

[36] S. Naik, S. Dhaneja, A. Khilari, S. Basu, A. Shetty, and C. Rodrigues, “Correlation of Trace detection in Gene Xpert MTB/RIF ultra with MGIT TB culture in a high TB-endemic Country,” Tuberculosis, vol. 154, p. 102666, Sep. 2025, doi: 10.1016/J.TUBE.2025.102666.

[37] N. J. Nantumbwe et al., “Deciphering rifampicin resistance among tuberculosis patients who have trace results on Xpert MTB/RIF ultra assay,” medRxiv, p. 2025.06.13.25329610, Jun. 2025, doi: 10.1101/2025.06.13.25329610.

[38] P. R. Torokaa et al., “The pattern of rpoB gene mutation of Mycobacterium tuberculosis and predictors of rifampicin resistance detected by GeneXpert MTB/RIF assay in Tanzania,” PLoS One, vol. 19, no. 8, Aug. 2024, doi: 10.1371/journal.pone.0296563.

[39] B. R. Mchaki, F. X. Mgaya, P. P. Kunambi, B. Hang’ombe, M. I. Matee, and M. Munyeme, “Comparative Performance of Line Probe Assay and GeneXpert in the Detection of Rifampicin Monoresistance in a TB-Endemic African Country,” Antibiotics (Basel), vol. 11, no. 11, Nov. 2022, doi: 10.3390/ANTIBIOTICS11111489.

[40] Q. Nie et al., “Phenotypic drug susceptibility characterization and clinical outcomes of tuberculosis strains with A-probe mutation by GeneXpert MTB/RIF,” BMC Infectious Diseases 2023 23:1, vol. 23, no. 1, pp. 832-, Nov. 2023, doi: 10.1186/S12879-023-08509-0.

